# The Melanoma Genomics Managing Your Risk Study randomised controlled trial: Statistical Analysis Plan

**DOI:** 10.1101/2020.04.13.20064246

**Authors:** Serigne N Lo, Amelia K Smit, David Espinoza, Anne E Cust, on behalf of the Managing Your Risk Study Group

## Abstract

**Background:** The Melanoma Genomics Managing Your Risk Study is a randomised controlled trial that aims to evaluate the efficacy of providing information on personal genomic risk of melanoma in reducing ultraviolet radiation (UV) exposure, stratified by traditional risk group (low or high phenotypic risk) in the general population. The primary outcome is objectively measured total daily Standard Erythemal Doses at 12 months. Secondary outcomes include UV exposure at specific time periods, self-reported sun protection and skin-examination behaviors, psychosocial outcomes, and ethical considerations surrounding offering genomic testing at a population level. A within-trial and modelled economic evaluation will be undertaken from an Australian health system perspective to assess the cost-effectiveness of the intervention.

**Objective:** To publish the pre-determined statistical analysis plan (SAP) before database lock and the start of analysis.

**Methods:** This SAP describes the data synthesis, analysis principles and statistical procedures for analysing the outcomes from this trial. The SAP was approved after closure of recruitment and before completion of patient follow-up. It outlines the planned primary analyses and a range of subgroup and sensitivity analyses. Health economic outcomes are not included in this plan but will be analysed separately. The SAP will be adhered to for the final data analysis of this trial to avoid potential analysis bias that may arise from knowledge of the outcome data.

**Results:** This SAP is consistent with best practice and will enable transparent reporting.

**Conclusion:** This SAP has been developed for the Melanoma Genomics Managing Your Risk Study and will be followed to ensure high-quality standards of internal validity and to minimise analysis bias.

**Trial registration:** Prospectively registered with the Australian New Zealand Clinical Trials Registry ACTRN12617000691347 (date registered: 15/05/2017).

## Introduction

### Preface

Melanoma is a serious form of skin cancer associated with significant morbidity and mortality but is highly preventable through individual behaviour change [1, 2]. Reduced sun exposure and improved primary prevention behaviours (i.e. sun protection) could prevent over 80% of melanoma diagnoses [3]. However, in high incidence countries such as Australia, preventive behaviours remain sub-optimal and there is a need to further improve melanoma prevention strategies aimed at the broader population [4].

Genomic risk information, based on multiple common genetic variants associated with elevated risk, could be integrated into improved melanoma prevention and early detection strategies [5, 6]. Social and health behaviour theory suggest that personal genomic risk information, alongside support and education on prevention and early detection, may be more effective in motivating behaviour change than ‘one size fits all’ standard approaches [7-9]. But the research evidence on genomic risk interventions in healthy participants is limited by small sample sizes and a high risk of bias, and few of these studies have focussed on melanoma prevention behaviours [10, 11].

The *Melanoma Genomics Managing Your Risk Study*, an innovative randomised controlled trial (RCT), aims to establish if providing personal genomic risk information modifies preventive behaviours, compared to receiving standard prevention advice on melanoma preventive behaviours. A pilot RCT demonstrating feasibility and acceptability, and the trial protocol, were previously published [12, 13]. The study was prospectively registered (ANCTRN 12615000356561). The trial completed the target accrual in February 2019, and all participants will be followed up for 12-months. The reporting of this statistical analysis plan (SAP) is in accordance with the guidelines for the content of SAPs in clinical trials [14].

### Purpose of the study and analyses

To evaluate the impact of personal genomic risk of melanoma information on motivating skin cancer prevention behaviours, and on broader social, psychological, ethical and economic outcomes.

## Study Aims and Outcome measures

### Aims

#### Primary

The primary aim is to evaluate the e□cacy of providing information on personal genomic risk of melanoma in reducing ultraviolet radiation (UV) exposure at 12-months, stratified by traditional risk group (low or high phenotypic risk) in the general population.

#### Secondary

The secondary aims are to evaluate:

1. the intervention’s e□ect at 12-months on
  a. time-specific UV exposure
  b. self-reported UV exposure
  c. sun protection behaviours
  d. skin examinations
2. the e□ect on other behavioural outcomes including:
  a. tanning
  b. sunburn frequency
  c. hypothesised mediators of behaviour change;
3. psychological outcomes, including skin cancer-related worry and distress;
4. the intervention’s e□ect on short-term outcomes at 1-month;
5. the impact of personal genomic risk categories (lower than average, average or higher than average) on primary and secondary outcomes in the intervention arm alone.
6. the experience of the intervention, psychological impact of undergoing genomic testing and receiving results, and communication with others about the results. Sub-studies beyond the scope of this SAP:
7. Qualitative sub-study assessing ethical considerations surrounding o□ering genomic testing at a population level, and psychosocial issues arising from the study processes that may a□ect wider implementation; and,
8. Cost-e□ectiveness analysis of the intervention at 12-months and longer term from an Australian health-system perspective (refer to Health Economics Analysis Plan (HEAP[15]) for more details),
9. Correlates of sun exposure, sun protection and skin examination behaviours, using baseline data.

### Outcome measures

#### Primary Outcome measure

The primary outcome for this study is total daily Standard Erythemal Doses (SEDs) measured 12-months after the baseline assessment (baseline questionnaire and UV dosimeter). A SED is an objective measure of UV exposure, measured using a time-stamped electronic dosimeter badge, mounted in light-weight custom-made wristbands attached to the left wrist (similar to wearing a watch) during daylight hours [16]. UV dosimetry is the gold standard for assessing personal UV exposure [17, 18]. UV dosimeters will be worn by all participants for 10 days at baseline, and again 12-months after baseline. A subgroup of ∼240 participants will also wear a UV dosimeter 1-month after receipt of the booklet/s. Total daily SEDs will be calculated as the weighted average of the average daily weekday and weekend SEDs (i.e. (weekday×5+weekend×2)/7). SED values will be log transformed for all analyses.

#### Secondary Outcome measures

**Time-specific UV exposure** captured from the time-stamped UV dosimeters. This will be calculated as the weighted average within given time-points: morning, peak and afternoon periods will be defined respectively as ‘6am-10am, 10am-2pm, and 2pm-8pm’ (without daylight savings).

**Self-reported UV exposure** on weekdays and weekends assessed using questionnaire items: *Thinking about the past month, please tell us the times of day as well as the usual length of time that you spent outside between 7am and 6pm on a typical weekday and weekend*. Length of time is collected in 15 minute intervals (0, <15, 15-29, 30-44, 45-60 minutes), and will be recoded to the mid-point of the respective exposure times. Total weekday and weekend sun exposure will be calculated as the sum of mid-point exposure times. Self-reported UV exposure will be calculated as the weighted average of the average weekday and weekend self-reported UV exposure (i.e. (weekday×5+weekend×2)/7).

**Sun protection behaviours** [19] assessed individually and summarised as a sun protection habits index, which is calculated as the mean of six protective behaviours scored on a 4-point Likert scale (1=never or rarely, 2=sometimes, 3=often, 4=always): *During the past month, when outside, how often did you*…

a. *wear sunscreen?*
b. *wear a shirt with sleeves that cover your shoulders?*
c. *wear a hat?*
d. *stay in the shade or under an umbrella?*
e. *wear sunglasses?*
f. *limit your time in the sun during midday hours?*

Sunscreen use is assessed using an item: *Have you used sunscreen in the past month?*

(Yes/No). If participants respond ‘Yes’ they are asked the following items:

- *Was this usually a high protection sunscreen (SPF 30 or more)?* (Yes/No).
- *How often on average have you used sunscreen in the past month?* (Less than one a week, 1-2 days a week, 3-5 days a week, 6-7 days a week)
- *On days that you have used sunscreen in the past month, how often did you apply it?* (free-text response, restricted to numbers)
- *Did you usually apply sunscreen (select one of the following): to all parts of your body exposed to the sun OR only to parts of your body that are prone to sunburn*.

Hat wear is assessed an additional item: *Please select the option which is most similar to your usual headwear when in the sun in the past month*. Response options are: no headwear, beanie, cap, legionnaires, bucket hat, wide brimmed, veil/burka.

**Whole-body skin examinations**, performed on oneself or by a partner or (in a separate question) by a health professional [19, 20] will be dichotomised as ‘yes’ vs. ‘no’. If response in the 12-month questionnaire to either:

a. *In the past 12 months, have you, your partner, friend or family member checked at least some of your skin for any suspicious spots that might be skin cancer?* or
b. *In the past 12 months, have you had a health professional (e*.*g. a doctor, specialist or nurse) check at least some of your skin for any suspicious spots that might be skin cancer?*

is ‘*yes’* then the extent of skin check (*1=All or nearly all of your body, 2=Part of your body, 3=Checking a specific mole or spot*) is assessed. If either self-check or health professional check respond ‘*All or nearly all of your body*’ then *Whole skin examinations* will be coded as 1: yes. All other responses (including those that have no skin checks) will be coded as 0: No.

**Intentional tanning frequency** will be measured on a 5-point Likert scale (1=never, 2=rarely, 3=sometimes; 4=often, 5=always) in response to the question: *In the past month, how often do you spend time in the sun in order to get a tan?*

**Sunburn frequency** over the previous month [20] will be collected as ‘0’, ‘1’, ‘2’, or ‘3 or more times’ in response to the question: *In the past month, how many times did you have a red or painful sunburn that lasted a day or more*? This will also be categorised as ‘any’ versus ‘none’.

**Hypothesised mediators of behaviour change** will be measured by the following items: Perceived severity of melanoma [21]; *To what extent do you agree or disagree with the following?*

- *I believe that melanoma is easy to cure*
- *I believe that melanoma can have very severe consequences*
- *Getting melanoma would be a big health threat for me*

A mean score will be derived from response options on a 5-point Likert scale: 1=strongly disagree, 2=disagree, 3=neither agree nor disagree, 4=agree, 5=strongly agree.

Perceived risk of melanoma [22];

*Write a number between 0 and 100 (where 0 means no chance and 100 means absolute certainty) to show what you think the chance is that you will develop melanoma during your remaining lifetime*.

*How confident are you in the number you wrote above?* Response options are on a 5-point Likert scale: 1=not at all confident, 2=slightly confident, 3=somewhat confident, 4=quite confident, 5 = very confident.[23]

*Compared to the average person of your sex and age, what do you think the chance is that you will develop melanoma in your lifetime?* Response options are on a 5-point Likert scale: 1=Much below average, 2=Somewhat below average, 3=Average, 4=Above average, 5=Much above average.

Confidence identifying melanoma) [21]; *How confident are you in your ability to identify melanoma?* Response options are on a 5-point Likert scale: 1=not at all confident, 2=slightly confident, 3=somewhat confident, 4=quite confident, 5 = very confident.

Perceived effectiveness of specific sun protection behaviours in reducing personal melanoma risk [24]; *For me, [using sunscreen/wearing protective clothing (such as long sleeves, long pants, hat)/avoiding sun exposure during midday hours] is (or would be) effective in reducing my risk of developing melanoma*. Response options for these three items are on a 5-point Likert scale: 1=not at all effective, 2=a little effective, 3=somewhat effective, 4=moderately effective, 5=very effective.

Perceived importance of protective behaviours; *How important are these activities?*

*Limiting sun exposure in the middle of the day; Wearing long-sleeved shirts; Wearing long pants; Wearing a wide-brimmed hat; Using sunscreen of SPF 30 or higher; Asking your doctor to conduct a skin examination; Checking your own skin for suspicious spots / moles; Staying in the shade*. Response options are on a 4 point Likert scale: 1=not at all important, 2=not very important, 3=somewhat important, 4=extremely important.

Perceived capability of performing protective behaviours; *How capable do you feel that you can do these things well, whether or not you intend to do them? (the same eight protective behaviours listed in e) above)*. Response options are on a 4 point Likert scale: 1=not at all capable, 2=not very capable, 3=somewhat capable, 4=extremely capable.

Social norms about skin examinations and sun protection [21]; *How many [adult members of your family/friends] check their own skin, or have their skin checked by someone else at least every 2 years?*; *How many of your [family members/friends] do you think regularly take precautions in the sun (such as using sunscreen, wearing protective clothing, avoiding sun during midday hours)?* Response options are: 1=None, 2=Some, 3=Half, 4=Most, 5=All, 6=I don’t know

Skin Self-Examination Attitude Scale (SSEAS) [25]; *To what extent do you agree with the following statements:*

- *It is important to check my skin for skin cancer even if I have no symptoms*
- *Checking my skin regularly is a priority for me*
- *I think I could find something suspicious on my skin if it was there*
- *If I saw something suspicious on my skin, I’d go to the doctor straight away*
- *I am confident in a doctor’s ability to diagnose skin cancer*
- *I am confident that I can take up examining my own skin even if I have not looked at my skin the past few months*
- *I am able to examine my own skin regularly, even if I have no one to help me*
- *If I regularly examine my skin, then I am helping to look after my own health*

Response options are on a 5-point Likert scale: 0=strongly disagree, 1=disagree, 2=unsure, 3=agree, 4=strongly agree. Responses to each item will be summed to generate a total score that can range between 0 and 40.

Tanning attitudes (Pro-tan score) [26]; *To what extent do you agree with the following statements:*

- *I feel more healthy with a suntan*
- *A suntan makes me feel better about myself*
- *A suntan makes me feel more attractive to others*
- *This summer I intend to sunbathe regularly to get a suntan*
- *Most of my close family think that a suntan is a good thing*
- *Most of my friends think a suntan is a good thing*
- *A suntan protects you against melanoma and other skin cancers*.

Response options are on a 5-point Likert scale: 1=strongly disagree, 2=disagree, 3=neither agree nor disagree, 4=agree, 5=strongly agree. Responses to the seven items will be summed, giving a total score between 7 and 35; those with a total score of 7–14 will be classified as ‘anti-tan’.

Anxiety about skin-examinations [25]; *I think checking my skin would make me anxious*. Response options are on a 5-point Likert scale: 1=strongly disagree, 2=disagree, 3=unsure, 4=agree, 5=strongly agree.

Perceived control over developing melanoma [24]; *Overall, how much personal control do you feel you have over [whether you develop melanoma in the future?*/ *detecting a future melanoma early in its development?]* Response options are on a 5-point Likert scale 1=no control, 2=not much control, 3=some control, 4=moderate control, 5=a lot of control.

Family communication; *In the past month, how frequently have you discussed with your family: using sunscreen; wearing protective clothing (such as long sleeves, long pants, hat); avoiding sun exposure during midday hours; getting a skin check by a health professional (e*.*g. GP or dermatologist); doing personal skin checks; spending time in the sun to get a tan*. Response options are on a 5-point Likert scale 1=not at all, 2=a little, 3=sometimes, 4=quite a bit, 5=a lot.

Vitamin D knowledge; *Do you believe that if you always protect all your skin from the sun, you are in danger of not getting enough Vitamin D?* Response options are: yes, no, I don’t know.

Health professional advice about sun protection and skin examinations; *In the past 12 months, has a health professional provided you with information or advice about sun protection, or how to check your skin for early signs of melanoma?* Response options are: yes, no.

Barriers to sun protection; *If you do not regularly use sunscreen, can you tell us the reason for this?; If you do not regularly wear protective clothing (such as long sleeves, long pants, hat), can you tell us the reason for this?; If you do not regularly limit your time in the sun during midday hours, can you tell us the reason for this?* Each of these items have a free-text response field and will be analysed using content and thematic analysis.

Other potential mediators include items on: the importance of personal health in achieving life goals [27] (assessed using a 5-point Likert scale); perceived general health [28] (assessed using a 5-point Likert scale); frequency of information-seeking related to skin cancer, genetics and genetic counselling; use of the study website; and use of applications (apps) to assist with managing sun protection or skin examinations.

### Psychological outcomes

Melanoma related worry will be assessed using three items:

i. *How worried are you about getting melanoma someday?*
ii. *How often does your worry affect your mood?*
iii. *How often does your worry affect your ability to perform your daily activities?*

Each item will be assessed on a 5-point Likert scale: 1=not at all, 2=rarely, 3=sometimes, 4=often, 5=almost all the time, and a mean score will be calculated.

Psychological distress and well-being will be assessed separately using the 5-item version of the Mental Health Inventory (MHI-5) [29]. The MHI-5 is a subscale of the SF-36 comprising the following items: *How much of the time during the past month have you:*

a. *been a very nervous person?*
b. *felt so down in the dumps that nothing could cheer you up?*
c. *felt calm and peaceful?*
d. *felt downhearted and blue?*
e. *been a happy person?*

Response options are on a 6-point Likert scale ranging from 1=never to 6=all of the time. Items c) and e) above will be reverse coded and the five scores then summed to a total subscale score in which higher scores indicate a worse health state and lower scores a better health state. Up to two missing values on the subscale will be imputed using the personal mean values from the completed items; otherwise the total score will be set to missing [30]. Each raw scale subscale score (ranging from 5-30) will be transformed to a 0 to 100 scale using the formula below, where the lowest possible raw score is 5 and the possible raw score range is 25: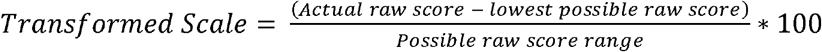

### Intervention arm additional measures

the experience of the intervention, psychological impact of undergoing genomic testing and receiving results the experience, and communication with others about the results, will be assessed using several items.

The psychological impact of undergoing testing and receiving personal genomic risk information will be assessed through the Multidimensional Impact of Cancer Risk Assessment (MICRA) [31]. The MICRA consists of three subscales a) distress (six items), b) uncertainty (nine items) and c) positive experiences (four items - reverse scaled).

**Table 1.**
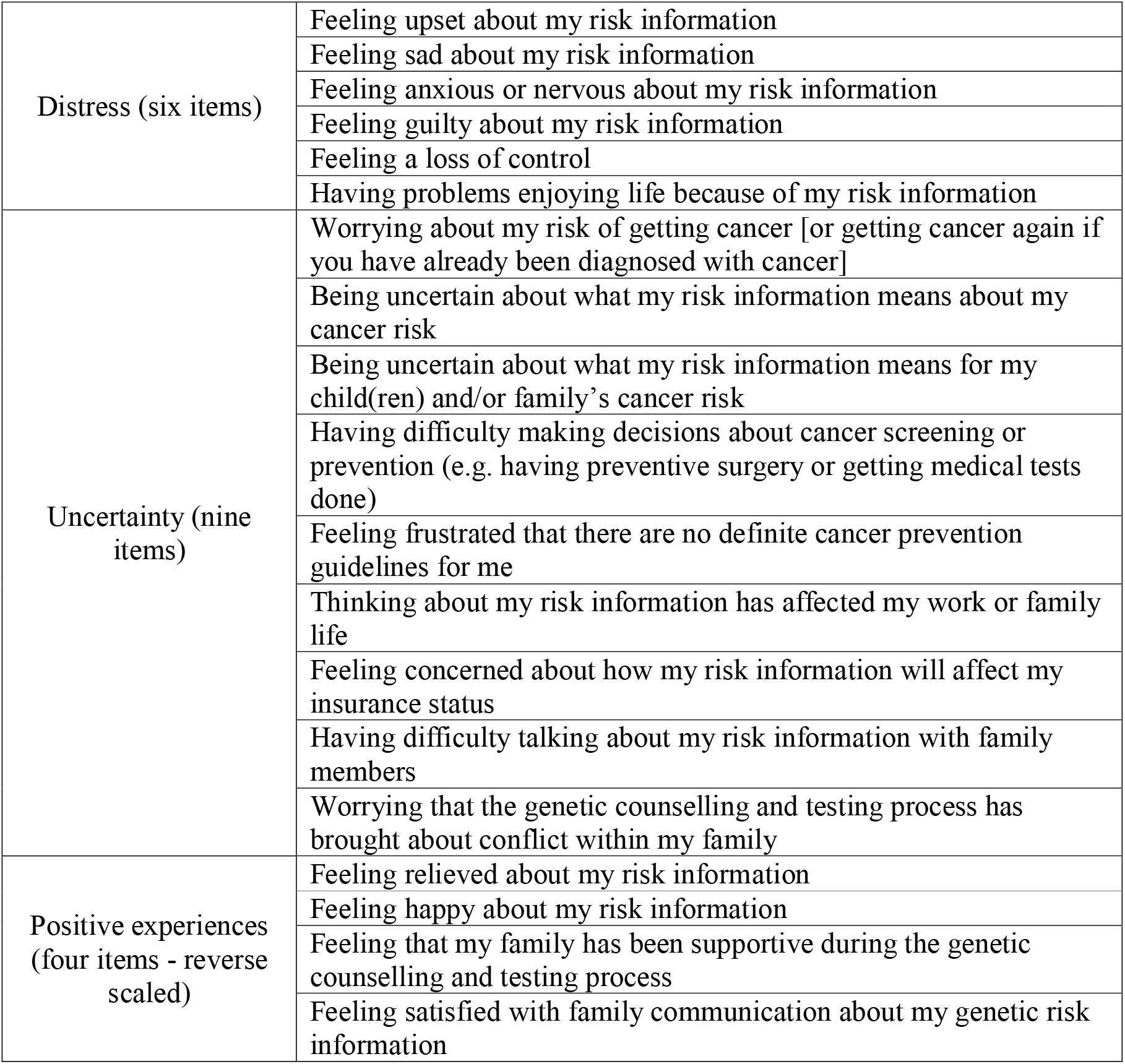
MICRA Domain Subscales.

Additional MICRA items that are assessed separately to the subscales include: Feeling regret about getting my risk information; Understanding clearly my choices for melanoma prevention or early detection. In addition, if the participant has a child(ren), two additional items are assessed: Worrying about the possibility of my children getting melanoma; Feeling guilty about possibly passing on the melanoma risk to my child(ren). If the participant has cancer, or has had it in the past, the following items are also assessed: Feeling that my genetic risk information has made it harder to cope with my cancer; Feeling that my genetic risk information has made it easier to cope with my cancer. The responses are scored as 0 for ‘never’, 1 for ‘rarely’, 3 for ‘sometimes’ and 5 for ‘often’ for the distress and uncertainty MICRA subscale items; and as 5 for ‘never’, 3 for ‘rarely’, 1 for ‘sometimes’ and 0 for ‘often’ for the positive experiences subscale items. The scores will be summed for each subscale, and the three subscales also summed to give a total score. Higher values of the MICRA score are related to increased experiences of distress. The questionnaire items for the additional MICRA items will be scored the same way as the distress and uncertainty subscales, with the possible range of 0-5 when there is one item, and 0-10 when there are two items.

Genetic counselling satisfaction survey (GCSS) [32]; *To what extent do you agree with the following statements?*

- *I felt I could talk about my reaction to my risk information with the genetic counsellor*
- *The genetic counsellor helped me to understand my risk information and make decisions about my health care*
- *I felt better about my health after talking to the genetic counsellor*
- *The length of the phone call was appropriate*
- *The genetic counsellor was truly concerned about my well being*
- *Talking to the genetic counsellor was valuable to me*
- *Understanding of genetic risk information, and amount read by participants*

Response options are on a 5-point Likert scale: 1=strongly disagree, 2=disagree somewhat, 3=uncertain, 4=agree somewhat, 5= agree strongly.

- Recall of personal genetic risk category
- Communication of result with family, friends and health professionals
- Motivations and barriers to family communication about genetic risk [33]
- Satisfaction, understanding and amount read of the personal genetic risk booklet

## Study Methods

### General Study Design and Plan

The Melanoma Genomics *Managing Your Risk Study* is a parallel group, two-arm, RCT with 1:1 allocation ratio. Supplementary Figure 1 shows the schema for the trial.

### Eligibility Criteria

Eligible participants met all of the following criteria:

1. Aged 18-69 years at the time of recruitment;
2. Never had a melanoma;
3. Some European ancestry, since current knowledge of genomic variation associated with melanoma risk is based predominately on populations with European ancestry, although there are ongoing efforts to improve this disparity [34].

### Randomisation and Blinding

Randomisation occurred once participants completed their baseline questionnaire and returned their baseline UV dosimeter. Randomisation to the intervention or control arm (allocation ratio 1:1) was conducted independently by the University of Sydney’s NHMRC Clinical Trials Centre Randomisation Service using a computer-based system. The stratified minimisation procedure [35] was used to ensure that the study groups were balanced by:

- traditional phenotypic risk score (low v high) [36],
- sex (male v female),
- state or territory of residence within Australia, and
- age group (18–44 years v 45–69 years).

Traditional risk scores were classified as high or low based on a validated published risk prediction model that includes variables for moles (naevi), hair colour, eye colour, artificial sunbed use, first-degree family history of melanoma, and personal history of keratinocyte cancers (e.g. basal cell carcinoma, squamous cell carcinoma) [36]. For the traditional risk score, a cut point of 1.223 was used to ensure approximately half of participants were in the low/high risk groups; this cut-point is based on data from our pilot trial and Australian Melanoma Family Study controls) [12, 36].

### Study Variables

Outcomes will be measured at baseline, 1-month after receipt of the booklet/s (except dosimeter outcomes that were only assessed for a subset of participants at 1-month), and again 12-months after baseline as summarised in 0. A sample of ∼240 participants will wear a UV dosimeter for 10-days at follow-up 1 (1 month after receipt of booklet/s).

**Table 2.** Schedule of study parameters and collection time points.

|  | STUDY PERIOD |  |  |  |  |
| --- | --- | --- | --- | --- | --- |
|  | Screening/<br>consent | Baseline/<br>randomisation | Intervention<br>delivery | Follow-up<sup>1</sup> |  |
| TIMEPOINT | -1 | 0 | T1 | F1 | F2 |
| **ENROLMENT:** |  |  |  |  |  |
| Eligibility screen | X |  |  |  |  |
| Informed consent | X |  |  |  |  |
| Allocation |  | X |  |  |  |
| **GROUP ALLOCATION:** |  |  |  |  |  |
| ***Intervention arm:*** |  |  |  |  |  |
| Saliva sample |  | X |  |  |  |
| Personalised melanoma genetic risk booklet |  |  | X |  |  |
| Phone call from genetic counsellor |  |  | X |  |  |
| Educational booklet on melanoma preventive behaviours |  |  | X |  |  |
| ***Control arm:*** |  |  |  |  |  |
| Educational booklet on melanoma preventive behaviours |  |  | X |  |  |
| **ASSESSMENTS** (control and intervention patients): |  |  |  |  |  |
| Demographics |  | X |  |  |  |
| Melanoma risk factors |  | X |  |  |  |
| Sun exposure (objective measure) |  | X |  | X<sup>2</sup> | X |
| Sun exposure (self-report) |  | X |  | X | X |
| Sun protection behaviours |  | X |  | X | X |
| Skin examination behaviours |  | X |  | X | X |
| Intentions, beliefs and attitudes towards sun protection behaviours and skin examinations |  | X |  | X | X |
| Discussion about sun protection and skin examinations with family |  | X |  | X | X |
| Perceived social norms about sun protection and skin examination |  | X |  |  |  |
| Perceived melanoma risk |  | X |  | X | X |
| Skin-cancer related worry |  | X |  | X | X |
| Perceived control over the development and early detection of future melanomas |  | X |  | X | X |
| Risk taking behaviours (Domain-specific risk-taking scale [DOSPERT]) |  | X |  |  |  |
| Psychological distress and well-being (MHI-5) |  | X |  | X | X |
| Confidence in completing medical forms |  | X |  |  |  |
| General health |  | X |  |  |  |
| Satisfaction with educational booklet on melanoma preventive behaviours, and amount |  |  |  | X | X |
| read by participants |  |  |  |  |  |
| Out-of-pocket costs for sun protection items |  |  |  | X | X |
| Visits to healthcare professionals |  |  |  | X | X |
| Private health insurance |  |  |  | X | X |
| Use of medications that may increase risk of melanoma or skin sensitivity to sunlight |  | X |  | X | X |
| Importance of health |  | X |  |  |  |
| Confidence in understanding medical information |  | X |  |  |  |
| Reasons for participating in the study |  | X |  |  |  |
| Receiving advice from health professional regarding sun protection and skin checks |  | X |  | X | X |
| Information seeking about skin cancer and genetics |  | X |  | X | X |
| Use of the study website |  | X |  | X | X |
| Use of applications related to sun protection or skin examinations |  |  |  |  | X |
| ***Measures related to receiving genetic risk information (intervention participants only):*** |  |  |  |  |  |
| Recalling personal genetic risk |  |  |  | X | X |
| Communication with family, friends and health professionals about genetic risk |  |  |  | X | X |
| Motivation+ barriers to communication about genetic risk |  |  |  | X | X |
| Satisfaction with genetic risk booklet and genetic counselling |  |  |  | X | X |
| Understanding of genetic risk information, and amount read by participants |  |  |  | X | X |
| Multidimensional impact of cancer risk assessment |  |  |  | X | X |

<sup>1</sup> Follow-up 1 (F1) takes place 1-month after participants receive their booklets. Follow-up 2 (F2) takes place 12-months after participants complete the baseline assessment (baseline questionnaire and UV dosimeter).
<sup>2</sup> The UV dosimeter measurement will be measured in ~240 participants rather than all participants at the short-term (1 month) follow-up.

### Sample Size

Sample size calculations are based on detecting a 20% difference in the primary outcome of average daily SEDs between intervention and control arms, in each of the low- and the high-risk phenotype groups. There is currently no consensus as to a ‘safe’ daily SED limit. Instead, skin cancer prevention strategies for the public primarily aim to shift the distribution of sun exposures downwards, especially in seasons of peak exposure. Due to the skewed nature of the SED exposure data, the sample size was calculated using a t-test with a geometric mean ratio (geometric mean SEDs in intervention group/geometric mean SEDs in control group) of 0.8, coefficient of variation 0.9, 80% power and α of 0.05.

Based on these calculations, a sample of 756 people (378 in the high-risk phenotype group and 378 in the low-risk phenotype group), split evenly between the intervention and control arms, will provide 80% power to detect a 20% reduction in the primary outcome in favour of the intervention with either low- or high-risk phenotypes. Allowing up to 15% with incomplete follow-up at 12 months, we will need to recruit 892 people (446 in each of intervention and control arms). Secondary analysis pooling the data from the low- and high-risk phenotype groups will give >97% power to detect a 20% difference in SEDs between the intervention and control groups.

No interim analysis, safety analysis and or data monitoring is planned for this study due to the minimal risks associated with the intervention.

### General Considerations for Statistical Analysis

The analysis principles are as follows:

- All analyses will be conducted on an intention-to treat basis. All randomised participants will be analysed in the group to which they were assigned.
- The primary analyses will stratify by phenotypic risk category (high, low).
- Statistical hypothesis tests will be evaluated at a nominal two-sided 5% level of significance.
- Intervention effect estimates (i.e. difference in means, odds ratio or relative risk) and their 95% confidence interval (CI) will be reported for all outcomes.
- The assessment of the overall intervention effect on outcome measures will be adjusted for baseline scores and randomised stratification factors.
- Subgroup analyses will be carried out irrespective of whether there is a significant effect of intervention on outcome.
- P values will not be adjusted for multiple comparisons.
- P values will be reported to three decimal places unless the p-value is less than 0.001 in which case it will be reported as ‘<0.001’. The mean, SD and any other statistics other than quantiles will be reported to one decimal place greater than the original data. Quantiles, such as median, or minimum and maximum will use the same number of decimal places as the original data. Estimated parameters, not on the same scale as raw observations (e.g. regression coefficients), will be reported to three significant figures.
- Analyses will be conducted primarily using SAS, version 9.4 or later and R 3.6.1 or later.

### Timing of Analyses

The final analysis will be performed after all randomised participants have completed their 12-month follow-up (or dropped out prior).

### Analysis Populations

The final analysis will include all participants who were randomised. The analysis population will be split into one of two phenotypic risk groups (i.e. high-risk and low-risk) as per the randomisation stratification variable ‘Traditional phenotypic risk score’. For secondary analysis of the primary outcome the analysis population will comprise the pooled phenotype risk groups.

### Covariates and Subgroups

The following subgroup analyses will be performed in each phenotypic risk groups and overall. It is hypothesized that the e□ect of the intervention on behaviour change or psycho-social outcomes may be influenced (moderated) by:

- Sex: male/other vs. female
- Age: 18–44 vs. 45–69 years
- State or territory of residence (based on latitude): Qld, NT, WA, NSW, ACT vs. SA, Vic, Tas
- Health literacy and numeracy: higher vs. lower [28]
- Family history of melanoma or a personal history of non-melanoma skin cancer: yes vs. no
- Education: school-only vs higher education
- Socio-Economic Indexes for Areas (SEIFA) using the Index of Relative Socio-Economic Advantage and Disadvantage (IRSAD) based on postcode of residence and categorised into 5 groups using quintile cut-points.
- Children: yes vs. no
- Risk taking propensity: risk-averse vs risk-seeking (see domain specific risk taking [DOSPERT] scale below [37]).
- Taking medications such as immuno-suppressants that may increase their sensitivity to sunlight (yes, no, don’t know).
- Genetic determinism, measured as *How much do you think genetic make-up, that is characteristics that are passed down from one generation to the next, determine whether or not a person will develop melanoma?* [38] on a 5-point Likert scale and categorised as 4,5 (completely/moderately) vs. 1,2,3 (not at all, slightly, somewhat).
- Genomic risk category (available for the intervention group only): higher than average vs. average vs. lower than average risk.
- Discordant genotype/phenotype groups (available for the intervention group only):
  - low-risk phenotype but high-risk genotype,
  - high-risk phenotype but low-risk genotype,
  - concordant combinations

### DOSPERT

The *Domain-Specific Risk-Taking (*DOSPERT) [37] scale contains three separate constructs: ‘risk-taking’, ‘risk-perceptions’, and ‘risk-attitude’ across separate domains. Two domains of risk, ‘Health and Safety’ and ‘Social’ were shown to be relevant to a preventive intervention for sun protection behaviours in our pilot study [13, 39] and were included in the baseline questionnaire; the items are shown in 03. For the assessment of:

- risk-taking, participants were asked to indicate ‘The likelihood you would engage in the described activity’, from extremely unlikely to extremely likely on a 7-point scale.
- risk perception, participants were asked to indicate ‘How risky you perceive the described situations’ from not at all risky to extremely risky on a 7-point scale,
- risk attitude, participants were asked to indicate ‘The benefits you would obtain from each situation’, from no benefits at all to great benefits.

**Table 3.** DOSPERT Domain Subscales.

| Domain Subscale | Item Text |
| --- | --- |
| Health/Safety | Drinking heavily at a social function. |
|  | Engaging in unprotected sex. |
|  | Driving a car without wearing a seat belt. |
|  | Riding a motorcycle without a helmet. |
|  | Sunbathing without sunscreen. |
|  | Walking home alone at night in an unsafe area of town. |
| Social | Admitting that your tastes are different from those of a friend. |
|  | Disagreeing with an authority figure on a major issue. |
|  | Choosing a career that you truly enjoy over a more prestigious one. |
|  | Speaking your mind about an unpopular issue in a meeting at work. |
|  | Moving to a city far away from your extended family. |
|  | Starting a new career in your mid-thirties. |

Item responses will be summed to obtain subscale scores for each domain for ‘Risk-Taking’, ‘Risk-Perceptions’, and ‘Risk-Attitude’. Individuals will be classified as ‘risk seeking’ if their subscale score was more than one standard deviation above the mean; ‘risk averse’ if their subscale score was more than one standard deviation below the mean; ‘risk neutral’ if their subscale score was within one standard deviation of the mean.

### Missing Data

The number of participants who complete the 12-month follow-up will be described by allocation; the study arms will be compared using chi-square tests and logistic regression to see if the attrition rate differs by arm and to compare baseline characteristics of participants who did and did not complete follow-up. Of those who complete follow-up, each variable will be examined for the presence of missing data and if >10% is observed for primary or secondary outcomes, then sensitivity analyses will be performed using complete case analysis or multiple imputation methods assuming data are missing at random (MAR). The MAR assumption indicates that the propensity for missingness does not depend on the unobserved outcome but rather is related to some other observed data.

The distribution of dosimeter sun exposure days will be described by study arm. Non-adherence will be defined as participants that fail to wear the dosimeter over at least one day on both a weekday and a weekend. Level of adherence (e.g. number of days worn) will also be described. For participants who do not have UV dosimeter (SED) data for both weekday and weekend exposures, we will estimate their missing exposure using imputation methods based on the participant’s available weekday or weekend SEDs, and SEDs data from the same age-group, gender, traditional (phenotypic) risk group and state or territory.

### Multiple Testing

There will be no formal adjustment for multiple comparisons. Outcomes are clearly categorised by degree of importance (primary and secondary) and cover a broad range of disciplines (e.g. behavioural, psychological, ethical, economic). Subgroup analyses have been pre-specified and are based on strong rationale and behaviour change theory.

### Summary of Study Data

Descriptive statistics will be prepared to summarise the data distributions and the characteristics of the study participants by allocation group. Continuous variables will be summarised using: n (non-missing sample size), mean (SD), median (25^th^-75^th^ centile) and categorical variables will be summarised by frequency and percentages (based on the non-missing sample size) of the observed levels.

### Subject Disposition

Subject disposition will be summarised with a CONSORT Flow Diagram (Supplementary Figure 2).

### Demographic and Baseline Variables

A list of baseline measures for all participants are shown in Supplementary Table 1.

### Intervention Compliance

We will document the number of people who completed the different aspects of the intervention, including providing a saliva sample, successful genotyping, receipt of booklets and phone call from the genetic counsellor.

### Efficacy Analyses

Efficacy analyses will be conducted on the basis of ‘intention to treat’. All group comparisons will be two tailed with a nominal 5% significance level, and will adjust for baseline scores and randomised stratification factors [40]. For (secondary) efficacy analyses that involve the pooled two phenotype risk groups, the model will also include phenotypic risk (high, low).

### Primary Efficacy Analysis

Geometric mean score and 95% CI will also be represented graphically from over time (baseline, follow-up 1 and 2). The primary analysis will be an intention-to-treat comparison of intervention and control arms for mean differences in UVR exposure measured as log-transformed daily SEDs at 12-months. The primary efficacy of the intervention will be assessed in each phenotypic risk group (high and low). UV dosimeter values will be log-transformed because of their right-skew distribution, and log-transformed values will be interpreted as a percentage change in the geometric mean of SEDs/day [41]. An Analysis of Covariance (ANCOVA), adjusted for baseline values and all randomised stratification factors will be performed [40]. A sensitivity analysis will be conducted on the subgroup of participants (N= ∼240) who will wear a UV dosimeter for 10-days at follow-up 1 (see section 0 for more details); given the repeated nature of the outcomes in this subgroup, generalized linear mixed models (GLMMs) with random intercepts will be fitted to assess the overall intervention effect.

### Secondary Efficacy Analyses

Where outcomes were assessed at three timepoints (baseline, 1-month, 12-months), including most questionnaire items, data will be analysed using GLMMs with random intercepts for continuous outcome measures, and generalised estimating equations (GEEs) with a log link function for binary outcome measures to estimate relative risks. The GLMM and GEE approach allows us to account for correlation due to repeated measures on each individual [42]. Where outcomes were assessed at two timepoints only (baseline and 12-months), data will be analysed using Analysis of Covariance (ANCOVA) for continuous outcome measures [40], and using GLM with log link function for binomial outcome variables to estimate relative risks and 95% CIs. The mean difference and 95% confidence intervals between intervention and control groups at 12-months will be estimated.

### Exploratory Efficacy Analyses

#### Subgroup analysis

The subgroup analyses will assess differences in intervention effects across pre-specified subgroups (refer to subheading ‘Covariates and subgroups’ pages 21-22). Tests of intervention effect modification will be performed by fitting intervention group and the relevant subgroup main effects and interaction into the models adjusted for baseline scores. Interpretation of evidence of heterogeneity of intervention effects among subgroups will remain exploratory (hypothesis generating) given the study is not powered to test subgroups in the stratified analysis. Results will be presented as forest plots with P values for heterogeneity (interaction test) for each pair of subgroups displayed.

### Sensitivity analysis

A sensitivity analysis of the repeated measures UV dosimeter data will be conducted on the subgroup of participants (N= ∼240) who wear a UV dosimeter at 1-month and 12-months, using GLMMs with random intercepts to account for the repeated measures.

A sensitivity analysis will also be completed comparing the results for people who completed follow-up in the Spring/Summer of 2018/19 vs. 2019/20 due to the bushfires experienced in Australia during the latter period and the potential impact on personal and ambient UV exposure.

A sensitivity analysis will be conducted in which dosimeter data will be used to adjust for the e□ects of measurement error in the relative risk estimate for self-reported sun exposure, using established methods [43].

### Technical Details

A second review statistician will independently reproduce the primary analyses. The reviewing statistician will have an overview of the entire analyses and will check the code, producing tables selected at random as well as any other pieces of code as desired.

## List of tables and figures

### Tables

1. Demographics and patient characteristics by intervention arm, stratified by phenotypic risk groups
2. Intervention compliance, stratified by phenotypic risk groups
3. Baseline characteristics by intervention arm, stratified by phenotypic risk groups
4. Data quality for primary and secondary endpoints, overall and by intervention arm
5. Intervention e□ct on 12-month UV exposure (SEDs), stratified by phenotypic risk groups (primary efficacy analysis)
6. The intervention’s effect on secondary outcomes (stratified by phenotypic risk groups):
  a. at 12-months on:
    ∘ time-specific UV exposure (SEDs)
    ∘ self-reported UV exposure
    ∘ sun protection behaviours
    ∘ skin examinations
  b. on other behavioural outcomes:
    ∘ tanning
    ∘ sunburn frequency
    ∘ hypothesised mediators of behaviour change
  c. on psychological outcomes, including skin cancer-related worry and psychological distress and well-being (MHI-5)
7. Intervention e□ect on primary and secondary outcomes overall
8. The intervention e□ect on short-term outcomes at 1-month (replicate Tables (5) and (6) for 1-month questionnaire measures and for the subgroup who completed a dosimeter at 1-month follow-up).
9. The impact of personal genomic risk category (low, average, high) on primary and secondary outcomes and receipt of genetic testing on outcomes in the intervention arm only
10. Subgroup analysis (replicate Tables (5) and (6) for each subgroup as defined in subgroups section)
11. Sensitivity Analysis
  a. Repeated measures analysis for participants who completed 1-month dosimeter measures
  b. The effect of time (Spring/Summer of 2018/19 vs. 2019/20) of intervention on UV radiation exposure (SEDs) and efficacy outcomes.
  c. Risk estimate for self-reported sun exposure adjusting for measurement error via use of dosimeter data.

### Figures

1. Trial flowchart (a Consolidated Standards of Reporting Trials [CONSORT]-style flow diagram to illustrate patient progression through the trial from initial screening for eligibility to completion of the final primary outcome assessment)
2. Forest plots of stratified and subgroup analyses.

## Data Availability

Not applicable.

---

**Supplementary Table 1.**
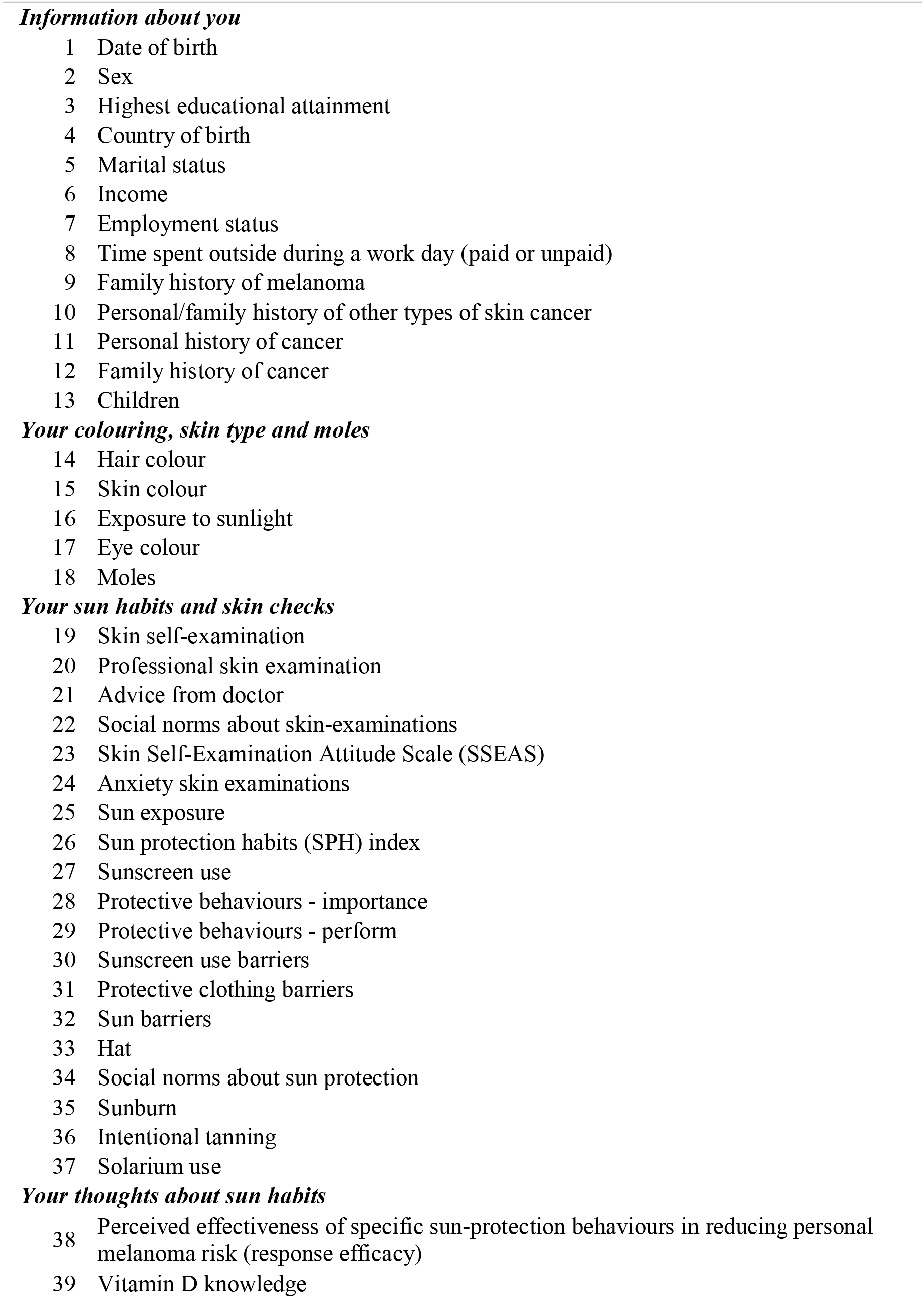
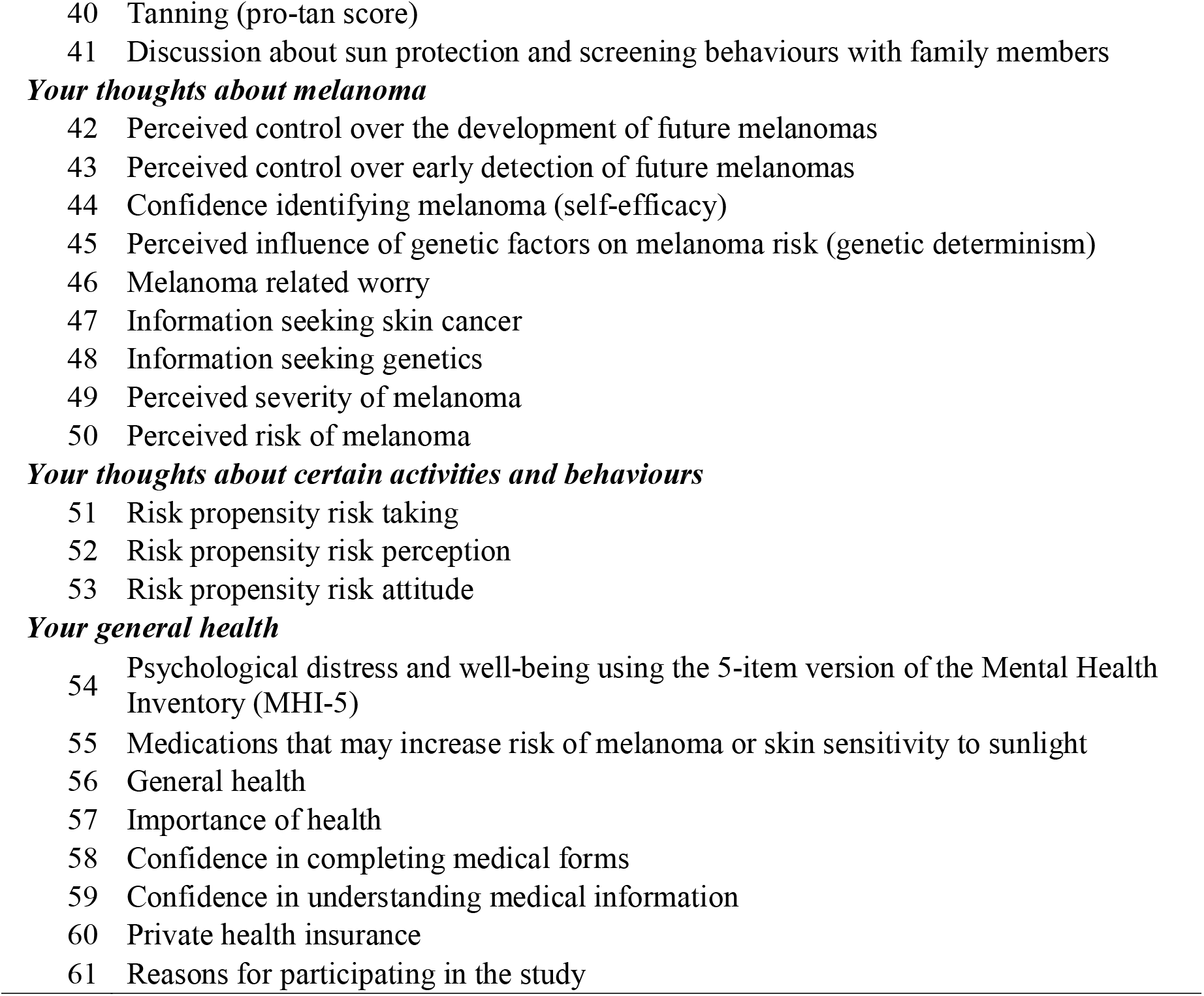
Demographic and Baseline Variables.

## Supplementary Figures

Supplementary Figure 1. Trial Schema

Supplementary Figure 2. CONSORT Flow diagram

## Abbreviations

CI: Confidence Interval
DOSPERT: Domain-Specific Risk-Taking
F1: Follow-up 1
F2: Follow-up 2
GEE: Generalised estimating equation
GLMM: Generalized linear mixed model
HEAP: Health Economics Analysis Plan
ICH E9: International Conference on Harmonisation, Statistical Principles for Clinical Trials
MAR: Missing At Random
MHI-5: Mental Health Inventory
MICRA: Multidimensional impact of cancer risk assessment
RCT: Randomised Controlled Trial
SAP: Statistical Analysis Plan
SD: Standard Deviation
SED: Standard Erythemal Dose
UV: Ultraviolet radiation.

## Ethics approval and consent to participate

Ethical approval has been obtained from the Human Research Ethics Committee at The University of Sydney (2017/163). The study was carried out in compliance with national and state legal and regulatory requirements and according to the International Principles of Good Clinical Practice (ICHGCP). As such all trial participants gave informed consent prior to joining the trial.

## Consent for publication

Not applicable.

## Availability of data and materials

Not applicable.

## Competing interests

None to declare

## Funding

This study is funded by the National Health and Medical Research Council of Australia (NHMRC; project grant 1129822). AEC receives a Career Development Fellowship from the NHMRC (1147843). AKS received a Research Training Program (RTP) Stipend Scholarship and a Merit Top Up Scholarship from the University of Sydney, and a Melanoma Institute Australia Postgraduate Research Scholarship. These funding bodies had no role in the design of the study and collection, analysis or interpretation of data or in writing the manuscript.

## Authors’ contributions

SNL, AKS, DE and AEC wrote and finalised the statistical analysis plan. All other authors provided input to the draft and read and approved the manuscript for publication. SL is the trial senior statistician, and AEC is the lead chief investigator for this trial.

## Acknowledgements

The study has been endorsed by the Melanoma and Skin Cancer Trials Group (ANZMTG 03.17). The authors and the authorship group would like to thank Bruce Armstrong for initial feedback on the study design, and Caro Badcock, Brooke Beswick, Georgina Fenton, Gillian Reyes-Marcelino, Ashleigh Sharman, Lauren Humphreys, Ainsley Furneaux-Bate, Rosa Evaquarta and Antigone Matsos for assistance with the conduct of this study.

## * Managing Your Risk Study Group

Anne E Cust^1,2^, Ainsley J Newson^3^, Rachael L Morton^4^, Michael Kimlin^5^, Louise Keogh^6^, Matthew Law^7^, Judy Kirk^8^, Suzanne J Dobbinson^9^, Peter Kanetsky^10^, Graham Mann^11^, Hugh Dawkins^12^, Jacqueline Savard^13^, Kate Dunlop^1^, Lyndal Trevena^1^, Mark Jenkins^14^, Martin Allen^15^, Phyllis Butow^16^, Sarah Wordsworth^17^, Serigne Lo^2^, Cynthia Low^18^, Amelia K Smit^1,2,3^, David Espinoza^4^

## Affiliations

^1^ The University of Sydney, Faculty of Medicine and Health, Sydney School of Public Health, Sydney, Australia

^2^ The University of Sydney, Melanoma Institute Australia, Sydney, Australia

^3^ The University of Sydney, Faculty of Medicine and Health, Sydney School of Public Health, Sydney Health Ethics, Australia

^4^ The University of Sydney, NHMRC Clinical Trials Centre, Australia

^5^ Cancer Council Queensland, Brisbane, Australia

^6^ Melbourne School of Population and Global Health, The University of Melbourne

^7^ Statistical Genetics, QIMR Berghofer Medical Research Institute, Brisbane, Australia

^8^ The University of Sydney, Faculty of Medicine and Health, Westmead Clinical School and Westmead Institute for Medical Research, Sydney, Australia

^9^ Cancer Council Victoria, Melbourne, Australia

^10^ H. Lee Moffitt Cancer Center and Research Institute and University of South Florida, U.S.

^11^ The John Curtin School of Medical Research, ANU College of Health and Medicine, The Australian National University, Canberra, Australia

^12^ HBF, Perth, Western Australia

^13^ School of Medicine, Faculty of Health, Deakin University, Geelong, Australia

^14^ Centre for Epidemiology and Biostatistics, Melbourne School of Population and Global Health, The University of Melbourne, Melbourne, Australia

^15^ Electrical and Computer Engineering, University of Canterbury, Christchurch, New Zealand

^16^ The University of Sydney, Faculty of Science, School of Psychology, Centre for Medical Psychology and Evidence-Based Decision-Making, Sydney, Australia

^17^ Health Economics Research Centre, The University of Oxford, Oxford, U.K.

^18^ Cancer Voices New South Wales, Sydney, Australia

## Notes

### Competing Interest Statement

The authors have declared no competing interest.

### Clinical Trial

Prospectively registered with the Australian New Zealand Clinical Trials Registry ACTRN12617000691347 (date registered: 15/05/2017).

## References

1. Green AC, Williams GM, Logan V, Strutton GM: Reduced melanoma after regular sunscreen use: randomized trial follow-up. J Clin Oncol 2011, 29:257–263.

2. Weinstock MA: Reducing death from melanoma and standards of evidence. J Invest Dermatol 2012, 132:1311–1312.

3. Armstrong BK, Kricker A: How much melanoma is caused by sun exposure? Melanoma Res 1993, 3:395–401.

4. Kasparian NA, McLoone JK, Meiser B: Skin cancer-related prevention and screening behaviors: a review of the literature. J Behav Med 2009, 32:406–428.

5. Burton H, Chowdhury S, Dent T, Hall A, Pashayan N, Pharoah P: Public health implications from COGS and potential for risk stratification and screening. Nat Genet 2013, 45:349–351.

6. Pashayan N, Hall A, Chowdhury S, Dent T, Pharoah PD, Burton H: Public health genomics and personalized prevention: lessons from the COGS project. J Intern Med 2013, 274:451–456.

7. Glanz K, Rimer BK, Viswanath K: Health Behavior. New York, United States: Wiley; 2014.

8. McBride CM, Birmingham WC, Kinney AY: Health psychology and translational genomic research: bringing innovation to cancer-related behavioral interventions. Am Psychol 2015, 70:91–104.

9. Rogers RW: A Protection Motivation Theory of Fear Appeals and Attitude Change. J Psychol 1975, 91:93–114.

10. Hollands GJ, French DP, Griffin SJ, Prevost AT, Sutton S, King S, Marteau TM: The impact of communicating genetic risks of disease on risk-reducing health behaviour: systematic review with meta-analysis. BMJ 2016, 352:i1102.

11. Marteau TM, French DP, Griffin SJ, Prevost AT, Sutton S, Watkinson C, Attwood S, Hollands GJ: Effects of communicating DNA-based disease risk estimates on risk-reducing behaviours. Cochrane Database Syst Rev 2010:CD007275.

12. Smit AK, Espinoza D, Newson AJ, Morton RL, Fenton G, Freeman L, Dunlop K, Butow PN, Law MH, Kimlin MG, et al: A pilot randomised controlled trial of the feasibility, acceptability and impact of giving information on personalised genomic risk of melanoma to the public. Cancer Epidemiol Biomarkers Prev 2016:10.1158/1055-9965.EPI-1116-0395.

13. Smit AK, Newson AJ, Morton RL, Kimlin M, Keogh L, Law MH, Kirk J, Dobbinson S, Kanetsky PA, Fenton G, et al: The melanoma genomics managing your risk study: A protocol for a randomized controlled trial evaluating the impact of personal genomic risk information on skin cancer prevention behaviors. Contemp Clin Trials 2018, 70:106–116.

14. Gamble C, Krishan A, Stocken D, Lewis S, Juszczak E, Dore C, Williamson PR, Altman DG, Montgomery A, Lim P, et al: Guidelines for the Content of Statistical Analysis Plans in Clinical Trials. JAMA 2017, 318:2337–2343.

15. Dritsaki M, Gray A, Petrou S, Dutton S, Lamb SE, Thorn JC: Current UK Practices on Health Economics Analysis Plans (HEAPs): Are We Using Heaps of Them? PharmacoEconomics 2018, 36:253–257.

16. Thieden E, Agren MS, Wulf HC: The wrist is a reliable body site for personal dosimetry of ultraviolet radiation. Photodermatol Photoimmunol Photomed 2000, 16:57–61.

17. Koster B, Sondergaard J, Nielsen JB, Allen M, Bjerregaard M, Olsen A, Bentzen J: Feasibility of smartphone diaries and personal dosimeters to quantitatively study exposure to ultraviolet radiation in a small national sample. Photodermatol Photoimmunol Photomed 2015, 31:252–260.

18. Dobbinson S, Niven P, Buller D, Allen M, Gies P, Warne C: Comparing Handheld Meters and Electronic Dosimeters for Measuring Ultraviolet Levels under Shade and in the Sun. Photochem Photobiol 2016, 92:208–214.

19. Glanz K, Yaroch AL, Dancel M, Saraiya M, Crane LA, Buller DB, Manne S, O’Riordan DL, Heckman CJ, Hay J, Robinson JK: Measures of sun exposure and sun protection practices for behavioral and epidemiologic research. Arch Dermatol 2008, 144:217–222.

20. Volkov A, Dobbinson S: 2010-11 National Sun Protection Survey: Report 2 prepared for National Skin Cancer Committee, Cancer Council Australia.

21. Kasparian NA, Branstrom R, Chang YM, Affleck P, Aspinwall LG, Tibben A, Azizi E, Baron-Epel O, Battistuzzi L, Bruno W, et al: Skin examination behavior: the role of melanoma history, skin type, psychosocial factors, and region of residence in determining clinical and self-conducted skin examination. Arch Dermatol 2012, 148:1142–1151.

22. Aspinwall LG, Taber JM, Kohlmann W, Leaf SL, Leachman SA: Perceived risk following melanoma genetic testing: a 2-year prospective study distinguishing subjective estimates from recall. J Genet Couns 2014, 23:421–437.

23. Cust AE, Armstrong BK, Smith BJ, Chau J, van der Ploeg HP, Bauman A: Self-reported confidence in recall as a predictor of validity and repeatability of physical activity questionnaire data. Epidemiology 2009, 20:433–441.

24. Aspinwall LG, Stump TK, Taber JM, Kohlmann W, Leaf SL, Leachman SA: Impact of melanoma genetic test reporting on perceived control over melanoma prevention. J Behav Med 2015, 38:754–765.

25. Djaja N, Youl P, Aitken J, Janda M: Evaluation of a skin self examination attitude scale using an item response theory model approach. Health Qual Life Outcomes 2014, 12:189.

26. Perez D, Kite J, Dunlop SM, Cust AE, Goumas C, Cotter T, Walsberger SC, Dessaix A, Bauman A: Exposure to the ‘Dark Side of Tanning’ skin cancer prevention mass media campaign and its association with tanning attitudes in New South Wales, Australia. Health Educ Res 2015, 30:336–346.

27. Marteau TM, Weinman J: Self-regulation and the behavioural response to DNA risk information: a theoretical analysis and framework for future research. Soc Sci Med 2006, 62:1360–1368.

28. Smith SK, Simpson JM, Trevena LJ, McCaffery KJ: Factors Associated with Informed Decisions and Participation in Bowel Cancer Screening among Adults with Lower Education and Literacy. Med Decis Making 2014, 34(6):756–72.

29. Berwick DM, Murphy JM, Goldman PA, Ware JE, Jr., Barsky AJ, Weinstein MC: Performance of a five-item mental health screening test. Med Care 1991, 29:169–176.

30. Peyre H, Leplege A, Coste J: Missing data methods for dealing with missing items in quality of life questionnaires. A comparison by simulation of personal mean score, full information maximum likelihood, multiple imputation, and hot deck techniques applied to the SF-36 in the French 2003 decennial health survey. Qual Life Res 2011, 20:287–300.

31. Cella D, Hughes C, Peterman A, Chang CH, Peshkin BN, Schwartz MD, Wenzel L, Lemke A, Marcus AC, Lerman C: A brief assessment of concerns associated with genetic testing for cancer: the Multidimensional Impact of Cancer Risk Assessment (MICRA) questionnaire. Health Psychol 2002, 21:564–572.

32. DeMarco TA, Peshkin BN, Mars BD, Tercyak KP: Patient satisfaction with cancer genetic counseling: a psychometric analysis of the Genetic Counseling Satisfaction Scale. J Genet Couns 2004, 13:293–304.

33. McGivern B, Everett J, Yager GG, Baumiller RC, Hafertepen A, Saal HM: Family communication about positive BRCA1 and BRCA2 genetic test results. Genet Med 2004, 6:503–509.

34. Bentley AR, Callier S, Rotimi CN: Diversity and inclusion in genomic research: why the uneven progress? J Community Genet 2017, 8:255–266.

35. Treasure T, MacRae KD: Minimisation: the platinum standard for trials?. Randomisation doesn’t guarantee similarity of groups; minimisation does. BMJ 1998, 317:362–363.

36. Vuong K, Armstrong BK, Weiderpass E, Lund E, Adami H, Veierod MB, Barrett JH, Davies JR, Bishop DT, Whiteman DC, et al: Development and external validation study of a melanoma risk prediction model based on self-assessed risk factors. JAMA Dermatol 2016, 152:9–6.

37. Blais AR, Weber EU: A Domain-Specific Risk-Taking (DOSPERT) scale for adult populations. Judgment and Decision Making Journal 2006, 1:33–47.

38. Hay J, Kaphingst KA, Baser R, Li Y, Hensley-Alford S, McBride CM: Skin cancer concerns and genetic risk information-seeking in primary care. Public Health Genomics 2012, 15:57–72.

39. Morton RL, Asher R, Peyton E, Tran A, Smit AK, Butow PN, Kimlin MG, Dobbinson SJ, Wordsworth S, Keogh L, Cust AE: Risk attitudes and sun protection behaviour: Can behaviour be altered by using a melanoma genomic risk intervention? Cancer Epidemiol 2019, 61:8–13.

40. European Medicines Agency Committee for Medicinal Products for Human Use (CHMP): Guideline on adjustment for baseline covariates in clinical trials. https://www.ema.europa.eu/en/adjustment-baseline-covariates-clinical-trials; 2015.

41. Vittinghoff E, Glidden DV, Shiboski SC, McCulloch CE: Regression Methods in Biostatistics. New York: Springer-Verlag; 2012.

42. Toshiro T: Repeated Measures Design with Generalized Linear Mixed Models for Randomized Controlled Trials. 1st edn: Chapman & Hall/CRC Biostatistics Series; 2017.

43. White E, Armstrong BK, Saracci R: Principles of Exposure Measurement in Epidemiology, Second Edition. New York: Oxford University Press; 2008.

